# Using an Ecological and Biological Framing for an Anti-racist Covid-19 Approach

**DOI:** 10.1101/2021.01.24.21250397

**Authors:** Araceli Camargo, Elahi Hossain, Sarah Aliko, Daniel Akinola-Odusola, Josh Artus, Ilan Kelman

## Abstract

In the United States and the United Kingdom COVID-19 has disproportionately affected Black, Indigenous and People of Colour (BIPOC) and Black, Asian and Minority Ethnic (BAME) people respectively. Multiple studies identify environmental factors such as overcrowded housing and poor workplace conditions as contributing factors for the disproportionate COVID-19 rates amongst BAME and BIPOC communities. This paper will show that to fully understand the phenomenon, both an ecological and biological approach is needed. An ecological approach highlights how a person’s habitat and the experiences within it mediate their susceptibility to disease. Moreover, to understand how this mediation works, this paper will use allostatic load as a biological pathway to link a person to their habitat and the poor health outcomes that contributed to COVID-19 susceptibility. In introducing this new approach, the paper will serve as an anti-racist framework for understanding how COVID-19 affected BAME and BIPOC communities. It is anti-racist by centring poor health outcomes on the habitats people are forced to live in due to structural racism rather than the physiology of a person’s race or ethnicity. This is important in order to avoid similar crises in the future and to improve the health of marginalised communities.

## Introduction

Globally, the COVID-19 pandemic has shone a light on deep and pervasive health inequities. There is statistical evidence both from the United States and the United Kingdom that COVID-19 mortality and infection rates have disproportionately affected Black, Asian, and Minority Ethnic (BAME)^1^ and Black, Indigenous, People of Colour (BIPOC) communities at higher rates.^2–4^ The coronavirus causing COVID-19, like most viruses, is a natural phenomenon; however, the deaths and their disproportionalities^2^ from the pandemic is by no means natural. In fact, it is a disaster,^5^ being “arising from a combination of hazard and vulnerability, with vulnerability as the causative factor”.^6^ Therefore, disasters, including this pandemic are not unavoidable, unpredictable or unprecedented.

Viewpoints of immediate and long-term vulnerability can be considered. From an immediate viewpoint, the disaster could have been avoided through following clear protocols. There are various examples of both local and international disease and pandemic strategies that could have been followed. The World Health Organisation (WHO) has various guides^7^ and national and local guidance also exists; for example, in the UK, simulations of future influenza pandemics have been conducted to understand how to execute effective UK specific responses^8^. Many governments, specifically those of wealthy nations like the UK and US failed to quickly heed the warnings from WHO. This combined with various other socio-political activities created vulnerabilities, resulting in ineffective responses.^6,9–11^

The next perspective to consider is the systemic vulnerabilities of public health, which include structural racism. Structural racism^12^ refers to instances of racial discrimination or inequity at an organisational or institutional level. For example, a country’s governance, which occurs through established rules, processes and opportunities, i.e. established policies, practices and norms which discriminate on the basis of race. This paper will contextualise how structural racism within the built environment and the places we inhabit plays a role generating disproportionate poor health outcomes in BAME and BIPOC communities. It has been well documented that BIPOC and BAME communities disproportionately inhabit neighbourhoods with higher levels of environmental pollutants, poor access to clean water, inadequate housing, and areas where there are food deserts.^13–19^ These environments, in turn, have potentially created vulnerabilities to COVID-19.

To explore this point further, a neurobiological lens has been introduced. The body and all its biological functions interact and link to the external world, in part through the stress response.^20^ Specifically, the stress response is there to react to stressors and keep biological function stable.^20^ Stress or a stressor is defined as an event that is perceived as a threat to biological stability. The brain classifies an event as a stressor as well as the physiological, cognitive and behavioural responses that follow.^21^ In order to adapt to the stressor, the brain activates a relay of responses through the activation of the Hypothalamic - Pituitary - Adrenal - Axis (HPA-Axis)^21,22^ and engages the endocrine, metabolic, and immune systems to afford necessary adaptations.^23^

Here lies the crucial link between current place habitation and vulnerability. As BAME and BIPOC communities disproportionately live in harsh environments, they are as a result exposed disproportionately to stressors, collectively called biological inequality.^24^ Biological inequality has previously been defined as a “comprehensive term that refers to the unequal distribution, exposure and vulnerability to health threatening pollution levels within urban environments”.^24^ Biological inequality contributes to a dysregulation of the stress response, which we see in people living in these communities. So, if environments or habitats play a key role in biological inequality and eventual poor health outcomes, then health can be seen as an ecological phenomenon, which is also directly in line with the WHO’s definition of health.^25^ More significantly, in order to remove vulnerabilities, an ecological approach to public health must be taken. This includes tackling social structures like racism which create one foundation of vulnerability, vulnerability-inducing policies, and mental frameworks of biological inequality. Finally, to create a clear link between habitat and person, the ecological case can be made through the use of neuroscience.

The term ‘inequality’ has typically been used to describe the difference in exposure to health-threatening levels of stressors across a population. However, due to structural racism an updated vocabulary is needed for accuracy. This paper adapts the term to become ‘biological inequity’. Linguistically, inequity captures the ‘unfairness’ of these differences, indicates the avoidability through better governance, and frames it as social injustice.^26^ This allows the phenomenon to be understood as a problem of structural racism. The term will also come to include psychosocial stressor exposure as a route to health-threatening biological differences. Indeed, psychosocial stressors in urban environments have been shown to contribute to health inequalities.^27,28^

Biological inequity can thus be defined as systematic, unfair, and avoidable stress-related biological differences which increase risk of disease, observed between social groups of a population. It can occur through structurally racist processes that result in disproportionate distribution and exposure to physical and psychosocial stressors. There are two main mechanisms by which social groups have an increased risk for disease from biological inequity: i) they are disproportionately exposed to the cause(s) of disease (differential exposure), and, ii) the effects of a cause(s) are different across groups (differential effects).^29^ For biological inequity, differential exposure is captured by the disproportionate distribution and exposure to physical and psychosocial stressors, while differential effects occur through stress-related biological differences brought about from the differential exposure.

First, this paper will detail the ecological link between people and health through the lens of neuroscience. Second, structural racism as the underpinning of this ecological link, and its consequent role in poor health outcomes, will be outlined. Third, it will use London, UK as a case study to illustrate the phenomenon using original data. Fourth, it will provide an anti-racist rehabilitation framework to help restore health in BAME and BIPOC communities.

### Ecological Approach and Cities

Ecology can be defined as the science and relationship between organisms and their environments.^30^ To effectively reduce vulnerabilities, which has led to communities being susceptible to COVID-19, an ecological approach to public health is needed, examining the interaction of multiple elements that affect a person’s health as part of a connected ecosystem which includes the places a person inhabits and their experiences within them.^30,31^ For example, when looking at obesity pathology, it is important to include environmental factors such as air pollution, given how it affects metabolic function.^32,33^

An ecological framing could encompass various geographies and scales; e.g., planetary, country, city, neighbourhood, or street. For this paper, a city habitat was chosen for three reasons. First, the ecosystem comprises living beings, natural biological ecosystems, micro climates, and biodiversity.^34^ Second, cities are becoming increasingly important habitats as more people move into them.^35^ Third, they are the most vulnerable habitats due to the consequences of structurally racist planning outcomes; climate inadequate city infrastructure, dilapidated transport systems, environmental pollutants, low biodiversity, and inadequate housing.^19,36–44^ The city of London in the U.K. was chosen due to accessible open source data, which facilitate the ability to analyse the distribution of environmental pollutants, inequities, and census data.

This type of approach allows for a systemic view of public health, specifically how the places we live play a role in long term health outcomes. However due to inequities, we have to look for who is being the most affected and who are the communities being forced to live in poor living conditions. Once this identification occurs, it is then possible to begin to understand how the planning of cities plays a role in the poor health outcomes of marginalised and discriminated communities. With the purpose of setting policies, recommendations, and health strategies which impede future inequities and reverse the outcomes of current ones. This is the anti-racist work that is needed to make cities healthier and more equitable.

### Structural Racism, Harsh Environments and Health

Structural racism in this case refers to the manner in which built environment factors, decisions, and policies are contributing to poor health outcomes in BAME and BIPOC communities.^45^ Structural racism, poverty and impoverished neighbourhoods are inextricably interlinked, with the intersection resulting in health inequities in urban environments. Through the systemic analysis of racism, studies in the US^46^ have shown that it is predominantly BIPOC communities that are forced to live in poorly planned and polluted environments. This phenomenon is also developing in the UK, where structural racism has shaped inequities in housing and proximity to pollution sources.^47^

These environments have been the source of many epidemiological studies, yielding a well substantiated theory identifying poverty as a central health risk.^48^ This then results in the health inequities seen, for instance, in that people living in low income households in the UK are two to three times more likely to have mental health difficulties.^49^ Thus, poverty is supported by racism, executed through urban systems, and experienced through poor health.

### Stress Response, Impoverished Urban Environments and Disease

The contribution of this paper is a proposal for the biological underpinnings of biological inequity. Specifically, the stress response and allostatic load play key roles in disease pathology. This paper considers two ecological factors: the physical environments people inhabit and the social experiences afforded by those environments. These factors are important for three reasons. First, both can be linked to the human biological system via the stress response. Second, these elements are highly influenced by structural racism. Third, understanding the role neighbourhoods and cities play in a person’s health outcomes provides an opportunity to look at the structural drivers. This is important for creating anti-racist health frameworks.

To recognise how the places people inhabit and their experiences shape, influence, and interact with human biology, an understanding of the stress response is necessary. The stress response is the main pathway linking the world around us with our health as it is a fundamental biological mechanism through which our bodies adapt to changing conditions (stimuli) in the environment that threaten biological stability.^20^ People encounter a variety of stressors categorised in two overarching types; internal and external. Internal stressors are generated within the body; for example, infections or thoughts. External stressors are generated outside the body; for example, major life events or pollution. Stressors have two further distinctions, physical and psychosocial. The physical categorisation includes environmental stressors, such as air, noise, light, or thermal pollutants.^50^ The psychosocial categorisation relates to experiences that occur within social environments, including social stressors, such as feeling socially isolated or experiencing discrimination (see Fig.1).

**Figure 1.**
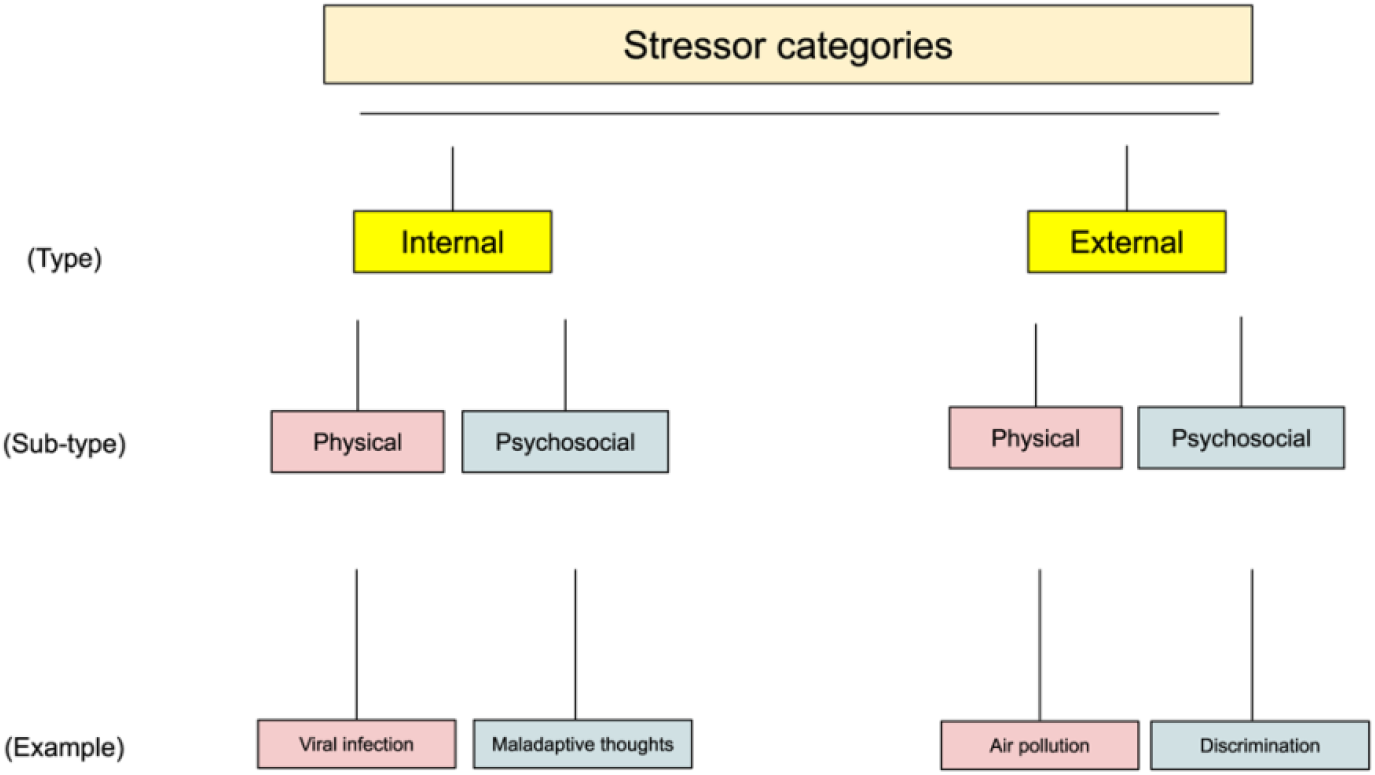
Stressor categories fall into two groups (internal and external), each of which can be either physical or psychosocial. The bottom row provides some real-world examples of stressors in each category.

Regardless of the stressor type, the body engages the stress response upon exposure. The stress response is mediated by the hypothalamic-pituitary-adrenal axis (HPA-Axis).^51^ In simple terms, its activation begins when a stressor triggers the hypothalamus, which is located in the brain and releases corticotropin releasing hormone (CRH).^52^ This trigger signals the pituitary gland, also located in the brain, to synthesise the adrenal-corticotropin releasing hormone (ACTH). ACTH is released into the circulation system where it reaches the adrenal glands, located above the kidneys.^52^ Once it reaches the adrenal glands, ACTH stimulates the release of cortisol, another regulatory hormone. Finally, cortisol circulates back to the brain through the circulatory system. Current understanding is that this cycle of hormones repeats in a negative feedback loop until the body reaches stability through various regulatory processes.^53^ This is called allostasis, a process by which “an organism maintains physiological stability by changing the parameters of its internal milieu, matching them appropriately to environmental demands”.^54^

The adaptation to stressors requires the HPA-Axis to interact with various other major systems such as endocrine, immune and metabolic systems. This interaction with the environment is long standing and is fundamental to how any biological system navigates and adapts with all aspects of their habitats.^54^ The stress response is in fact an integral part of adaptation and human survival.^55^ However, when the system is chronically engaged due to persistent exposure to stressors, the HPA-axis can become dysregulated and prevent the body from achieving optimal allostasis, leading to what is called allostatic load (AL).

AL represents the “wear and tear” of bodily systems caused by stressors from an individual’s environment.^56,57^ Over time, allostatic load impairs the function of various biological systems including immune, endocrine, digestive, and metabolic which can eventually lead to disease.^58,59^ This systemic and vast biological degradation is at the core of disease pathology. Diseases being both mental disorders such as depression and anxiety and physical disorders such as diabetes and obesity. It is important to note that AL is an established concept in neuroscience and epidemiology,^60^ making it a robust and reliable biological mechanism from which to approach the ecology of health.

AL needs to be contextualised to the habitats BAME and BIPOC communities are forced to live in due to poverty, which is a consequence of structural racism. BAME and BIPOC communities experience poverty at a disproportionate rate, which puts them at higher risk for biological inequity, resulting in disease susceptibility.^61–63^ Table 1 shows the different stressors communities experience in harsh environments due to structurally racist driven poverty. They are categories in the two main ecological pathways; the physical stressors present in these environments, as well as the psychosocial stressors from the lived experience of these environments. In many cases a stressor can be a mixture of psychosocial and physical. Indeed, these terms can be conceptualised as forming the opposing ends of a spectrum.^64^ For example, air pollution can be inhaled and impact physical systems and organs. Inversely, if a person is aware that they are experiencing air pollution and perceives it as a danger (threat) which they have no control over, they may also be affected at a psychosocial level. Therefore, it must be noted that the tick is the dominant categorisation rather than the sole categorisation.

**Table 1.**
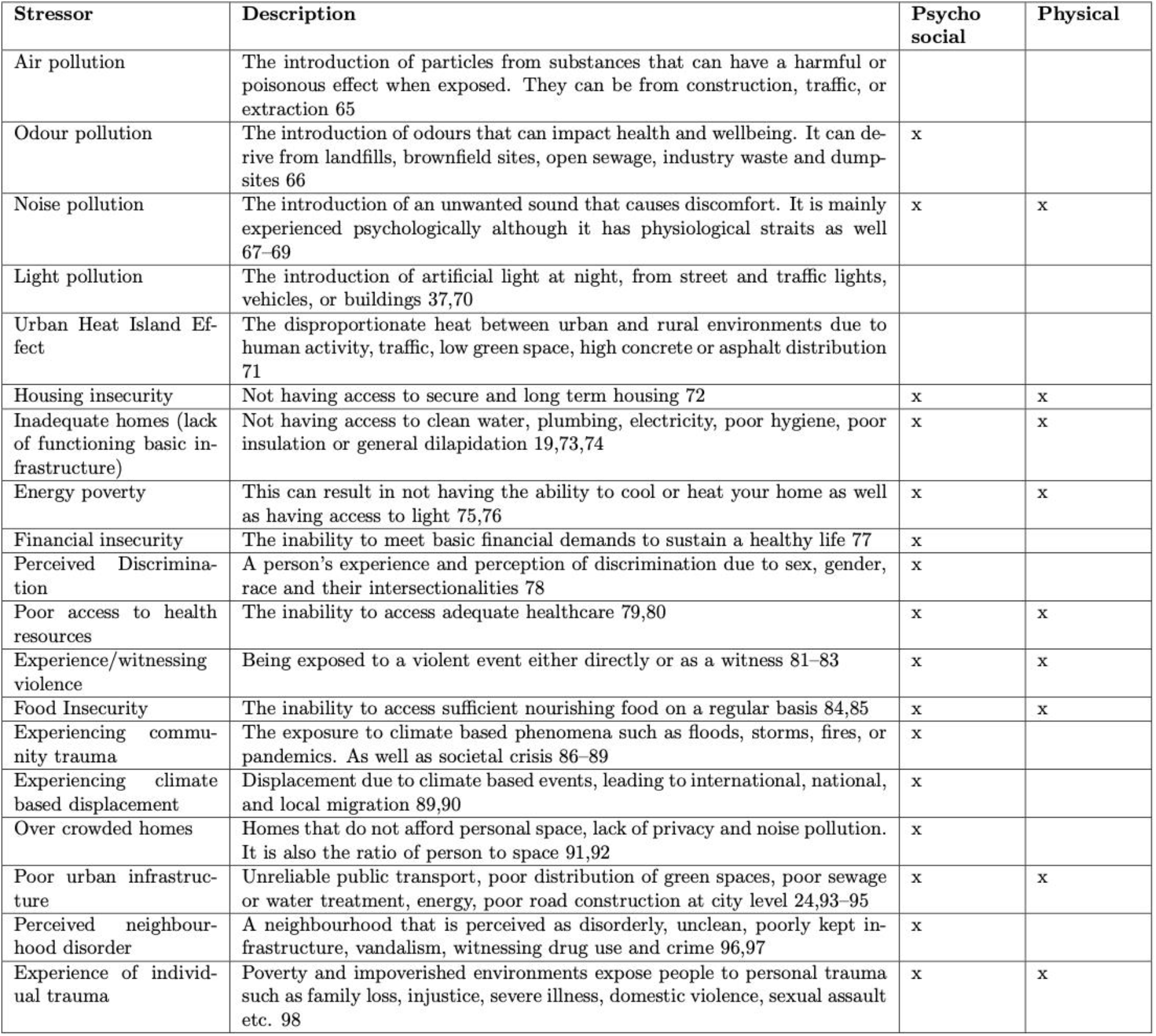
List of ecological stressors that are disproportionately experienced by BIPOC and BAME communities due to structural racism. Here we describe each type of stressor as defined by the literature and categorise them by psychosocial vs physical type of stressor.

| Stressor | Description | Psycho social | Physical |
| --- | --- | --- | --- |
| Air pollution | The introduction of particles from substances that can have a harmful or poisonous effect when exposed. They can be from construction, traffic, or extraction 65 |  |  |
| Odour pollution | The introduction of odours that can impact health and wellbeing. It can derive from landfills, brownfield sites, open sewage, industry waste and dumpsites 66 | x |  |
| Noise pollution | The introduction of an unwanted sound that causes discomfort. It is mainly experienced psychologically although it has physiological strains as well 67–69 | x | x |
| Light pollution | The introduction of artificial light at night, from street and traffic lights, vehicles, or buildings 37,70 |  |  |
| Urban Heat Island Effect | The disproportionate heat between urban and rural environments due to human activity, traffic, low green space, high concrete or asphalt distribution 71 |  |  |
| Housing insecurity | Not having access to secure and long term housing 72 | x | x |
| Inadequate homes (lack of functioning basic infrastructure) | Not having access to clean water, plumbing, electricity, poor hygiene, poor insulation or general dilapidation 19,73,74 | x | x |
| Energy poverty | This can result in not having the ability to cool or heat your home as well as having access to light 75,76 | x | x |
| Financial insecurity | The inability to meet basic financial demands to sustain a healthy life 77 | x |  |
| Perceived Discrimination | A person's experience and perception of discrimination due to sex, gender, race and their intersectionalities 78 | x |  |
| Poor access to health resources | The inability to access adequate healthcare 79,80 | x | x |
| Experience/witnessing violence | Being exposed to a violent event either directly or as a witness 81–83 | x | x |
| Food Insecurity | The inability to access sufficient nourishing food on a regular basis 84,85 | x | x |
| Experiencing community trauma | The exposure to climate based phenomena such as floods, storms, fires, or pandemics. As well as societal crisis 86–89 | x |  |
| Experiencing climate based displacement | Displacement due to climate based events, leading to international, national, and local migration 89,90 | x |  |
| Over crowded homes | Homes that do not afford personal space, lack of privacy and noise pollution. It is also the ratio of person to space 91,92 | x |  |
| Poor urban infrastructure | Unreliable public transport, poor distribution of green spaces, poor sewage or water treatment, energy, poor road construction at city level 24,93–95 | x | x |
| Perceived neighbourhood disorder | A neighbourhood that is perceived as disorderly, unclean, poorly kept infrastructure, vandalism, witnessing drug use and crime 96,97 | x |  |
| Experience of individual trauma | Poverty and impoverished environments expose people to personal trauma such as family loss, injustice, severe illness, domestic violence, sexual assault etc. 98 | x | x |

Given how many different factors in a person’s habitat and their lived experience affect health, the ecological perspective is needed for understanding how elements of the physical and social environment interact at multiple levels and for informing more effective urban policy interventions.^99^

### Susceptibility and COVID-19

The persistent and disproportionate exposure to health threatening levels of environmental and psychosocial stressors has been identified and defined as ‘biological inequity’.^24^ This puts a burden on the stress response, contributing to its dysregulation. Over time, this dysregulation impacts immune, metabolic, and endocrine systems, increasing an individual’s susceptibility to a wide range of diseases.^100^

Susceptibility refers to both the heightened risk of developing diseases and suffering more acute symptomatology compared to a normative population. There are two pathways to COVID-19 susceptibility (see Fig.2).^101^ The first is through biological inequity induced disease. Higher levels of biological inequity are associated with the development of diabetes, asthma, and obesity through allostatic load processes ^102^. These biological changes can heighten the risk of viral contraction, severity of symptoms, and mortality in relation to COVID-19.^103,104^ The second pathway is directly through allostatic load. A person may not seemingly have an underlying diagnosed disorder, but due to living in biological inequity they experience higher levels of chronic stress, i.e. allostatic load, which dysregulates the immune response.^105^ For example, in a study on respiratory infections, the “rates increased in a dose-response manner with increases in the degree of psychological stress”.^106^

**Figure 2.**
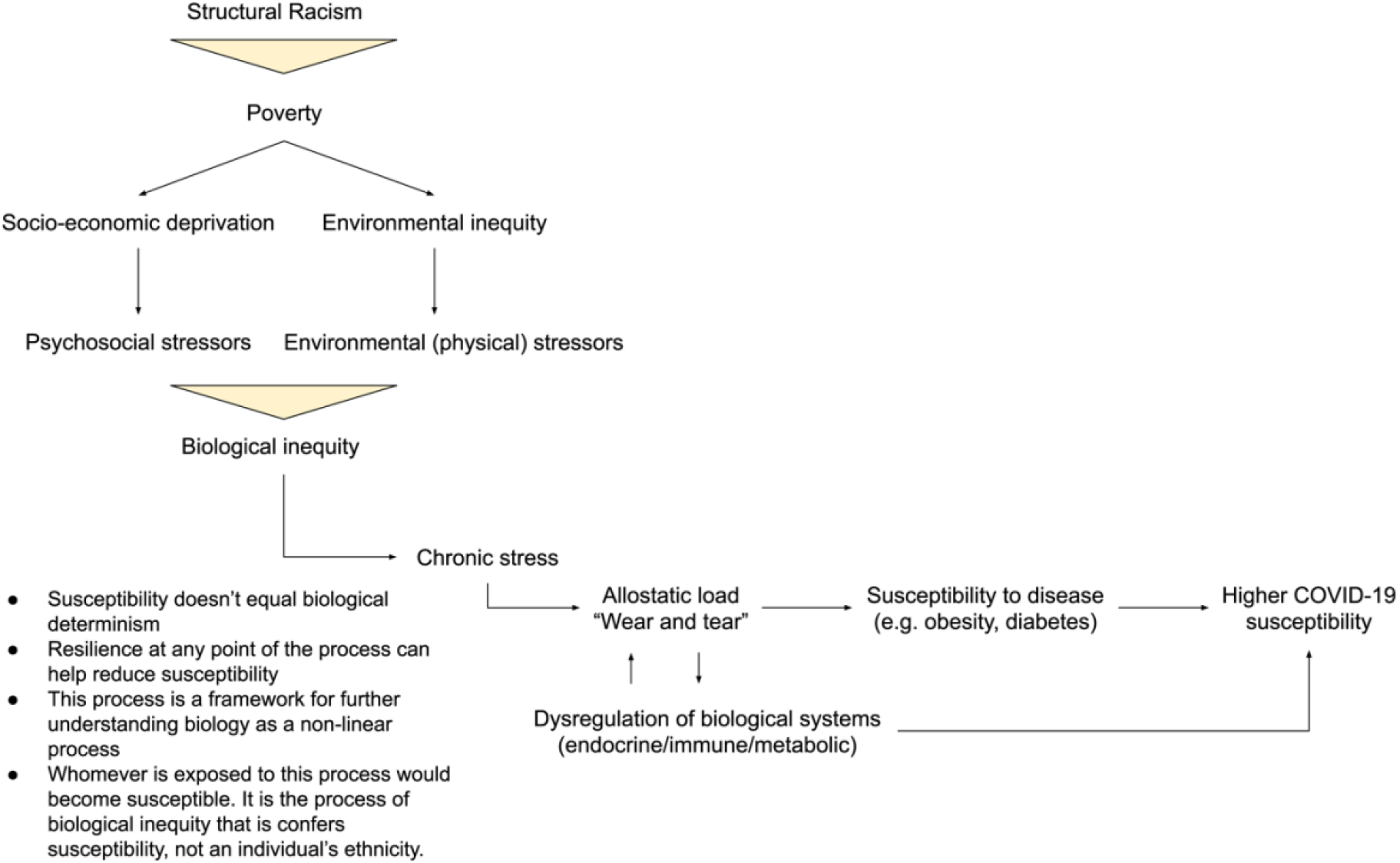
Pathway to higher COVID-19 susceptibility. Structural racism and biological inequity lead to chronic experience of stress, causing a ‘wear and tear’ on the immune system. The dysregulated allostatic load and immune system increase susceptibility to disease.

Two caveats should be highlighted. First, susceptibility is not the same as determinism. Being susceptible to disease does not mean that a person will certainly develop a disease as many other factors contribute to disease pathology. For example, a study in the US indicated that having emotional support can offset some of the effects of neuroendocrine, metabolic, inflammatory, profiles that tend to develop after exposure to acute and persistent stress.^107^ The second caveat is that it is not a matter of being BAME or BIPOC that is a health risk; it is the conditions and environments in which BAME and BIPOC communities are forced to live--namely racism and inequity, that is the health risk. This also covers the jobs they might have (e.g. bus drivers) which might put them in contact with more people; that they might be disproportionately forced to go to work on public transport due to where they live or lack of access to alternatives; and that they might have had fewer options regarding working from home or not working. These factors are relevant for the materials and methods, but we are focusing on the biological outcomes, not the possible difference in exposure to viral load or to more people on a daily basis.

## Materials and Methods

London is an international city with a diverse population of around 9 million^108^ comprising f 32 boroughs and the City of London^109^ with open access to data needed for mapping ecological health analyses and investigating structural racism.

In order to map the race-related susceptibility to COVID-19 within London, this study analysed four data sets: Index of Multiple Deprivation, Stress Risk Score, Census data and COVID-19 mortality rates obtained from the Office of National Statistics.

The Index of Multiple Deprivation (IMD)^110^ is the official measure of relative deprivation in England and is part of a suite of outputs that form the Indices of Deprivation (IoD). It follows an established methodological framework in broadly defining deprivation so that it encompasses a wide range of an individual’s living conditions:

Income. (22.5%)

Employment. (22.5%)

Education. (13.5%)

Health. (13.5%)

Crime. (9.3%)

Barriers to Housing and Services. (9.3%)

Living Environment. (9.3%)

People may be considered to be living in poverty if they lack the financial resources to meet their needs, whereas people can be regarded as deprived if they lack any kind of resources, not just income. Due to the robustness of the IMD^16^, it was used as a proxy for psychosocial stressors related to living in areas of deprivation.

The Stress Risk Score (SRS)^24^ is a scale used to measure the environmental stress risk factors based on proxies for noise, air, light, and thermal pollution, using a meta-analysis of how each stressor engages with the stress response, specifically allostatic load. The SRS is based on a cardinal linear scale of 0 to 4, with 0 being a less polluted area and 4 being highly polluted across four environmental stressors. The total score is obtained by summing the means of the individual pollutant scores. These individual scores are divided into bins from 0 to 1 based on thresholds from past neuroscientific research on pollutant effects on health that are mediated through stress-related pathways. The thresholds are maximum values that the pollutant can reach before human health is significantly at risk. The SRS was used to look at environmental stressors.

The Office of National Statistics (ONS)^111^ database provided data on age-standardised mortality rates (ASMR) for deaths involving COVID-19, local authorities in England and Wales and the 2011 England and Wales Census on BAME population density per 100,000. From the ONS database, only data concerning local authorities in London were included in the analysis.

BAME population density per 100,000 from the ONS database was provided at the borough level. Data from City of London was not included in the analysis due to inconsistencies in reporting across different datasets. In order to align all data for analysis, all other datasets were aggregated up to the borough level if they were at a higher spatial resolution (i.e. wards, postcodes etc).

The SRS scores, calculated at the individual Lower Super Output Area (LSOA) level, were summarised over each borough. LSOAs are the smallest geographical hierarchy available for London.^112^ In order to consider the difference in size of each LSOA in a borough, the SRS was weighted using hectare area of each LSOA, following a standard normalisation approach:

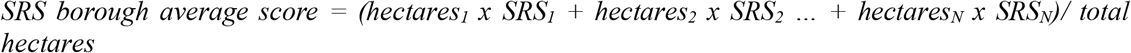

Although we acknowledge that wind, temperature and other meteorological factors help predict pollution spread over an area, the data is not available for the needed locations and scale. Thus, the weighted SRS average takes surface area into consideration, making it the best estimate for the data available.

The IMD average scores at the borough level were already provided by the ONS database, calculated using the same weighted method as the SRS, but with population size.

Pearson’s correlation was used to investigate the relationship between one of the scores and age-standardised COVID-19 mortality rates, as well as between BAME population density and age-standardised COVID-19 mortality. No causal relationship can be inferred from the data, thus correlation is applied to estimate the strength and direction of the relationship between any two linear variables. A linear regression between age-standardised COVID-19 mortality and BAME density per 100,000 population was computed.

Finally, the scores, BAME density and age-standardised COVID-19 mortality rates were overlaid onto the London borough map, in order to qualitatively inspect boroughs with high structural racism.

## Results

The data indicates high correlation between COVID-19 deaths and IMD 2019 by borough (r = 0.71) and between COVID-19 deaths and BAME population density (r = 0.82) during the peak of the first COVID-19 wave in the UK (see Table 2 March-April 2020). Moreover, a high correlation was observed between BAME density and IMD 2019 with COVID-19 deaths at the end of the first wave in July 2020 (r = 0.81 and r = 0.65 respectively). The SRS score had a medium correlation (r = 0.42, p < 0.05) with COVID-19 deaths during the April peak, but no significant correlation by the end of the first wave. Moreover, the SRS score and BAME population density had a medium-low positive correlation (r = 0.36, p < 0.05), while IMD with BAME population density had a higher positive correlation (r = 0.57, p < 0.05). Since SRS and IMD are highly correlated (r = 0.52), we will only address IMD from now on, as it encompasses both environmental and psychosocial stressors.

**Table 2.** Summary of Pearson’s correlation r values between age-standardised COVID-19 deaths per 100k population, BAME density per 100k population, SRS and IMD scores across London. There was a significant positive correlation between IMD (deprivation index) and BAME density and age-standardised COVID-19 deaths. No significant correlation was found between SRS scores and COVID-19 deaths.

| r value | p value <0.05 | Variable 1 | Variable 2 |
| --- | --- | --- | --- |
| 0.71 | Yes | IMD 2019 average scores by borough | Age-standardised COVID-19 deaths for 100k population (March-April 2020) |
| 0.42 | Yes | SRS average scores by borough |  |
| 0.82 | Yes | BAME density for 100k population |  |
| 0.65 | Yes | IMD 2019 average scores by borough | Age-standardised COVID-19 deaths for 100k population (March-July 2020) |
| 0.25 | No | SRS average scores by borough |  |
| 0.81 | Yes | BAME density for 100k population |  |

The direction and strength of the relationship between the age-standardised COVID-19 mortality and BAME density per 100,000 population were further explored qualitatively and summarised in Fig.3-4. Fig.3 shows a positive relationship between (normalised) COVID-19 mortality and BAME density across boroughs. For instance, Newham and Brent experienced the highest COVID-19 mortality and also have the highest BAME population density.

**Figure 3.**
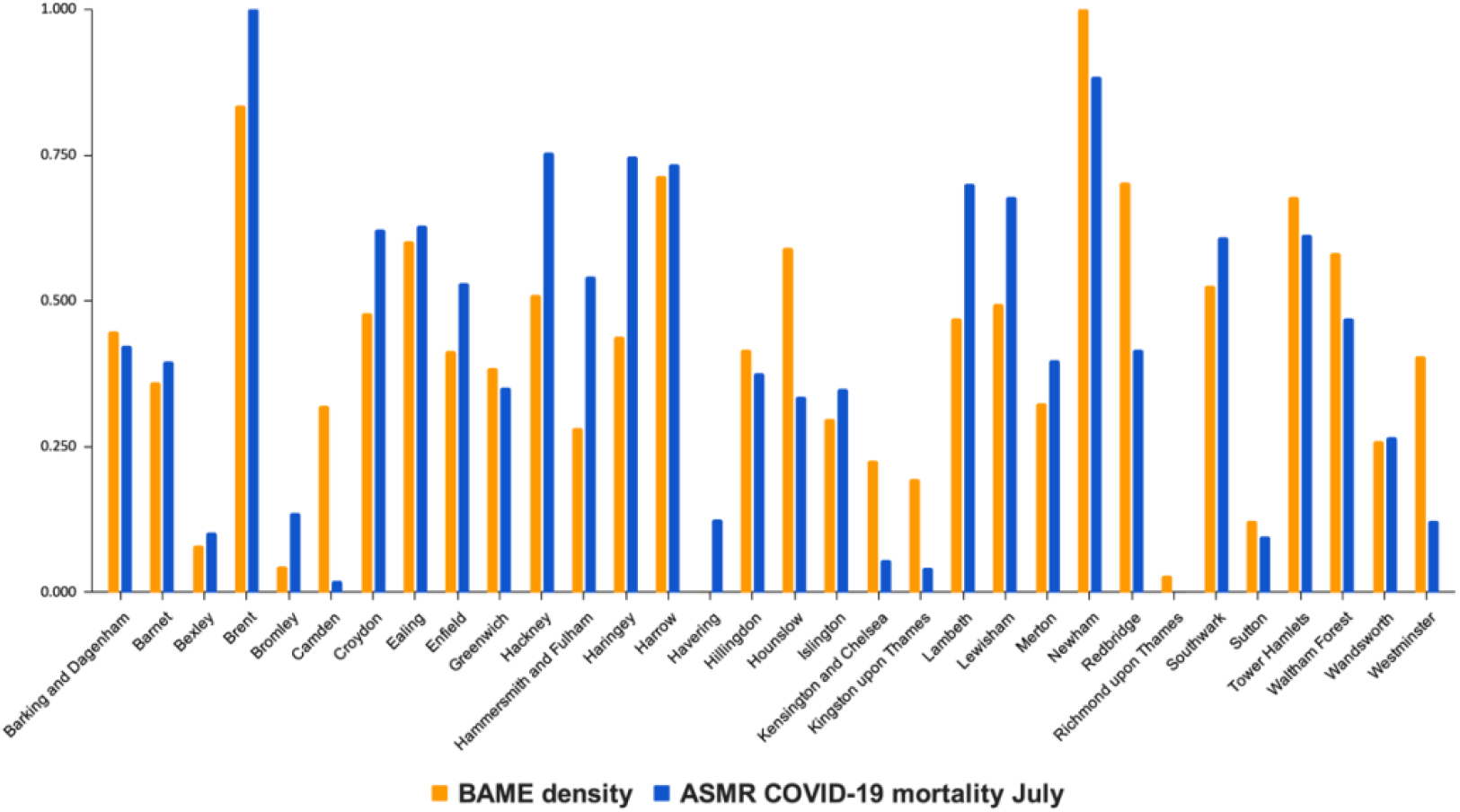
Relationship between normalised age-standardised COVID-19 mortality rates and normalised BAME density per 100k population across 32 boroughs in London, showing a positive correlation. The boroughs of Brent and Newham indicate highest BAME density and COVID-19 deaths.

**Figure 4.**
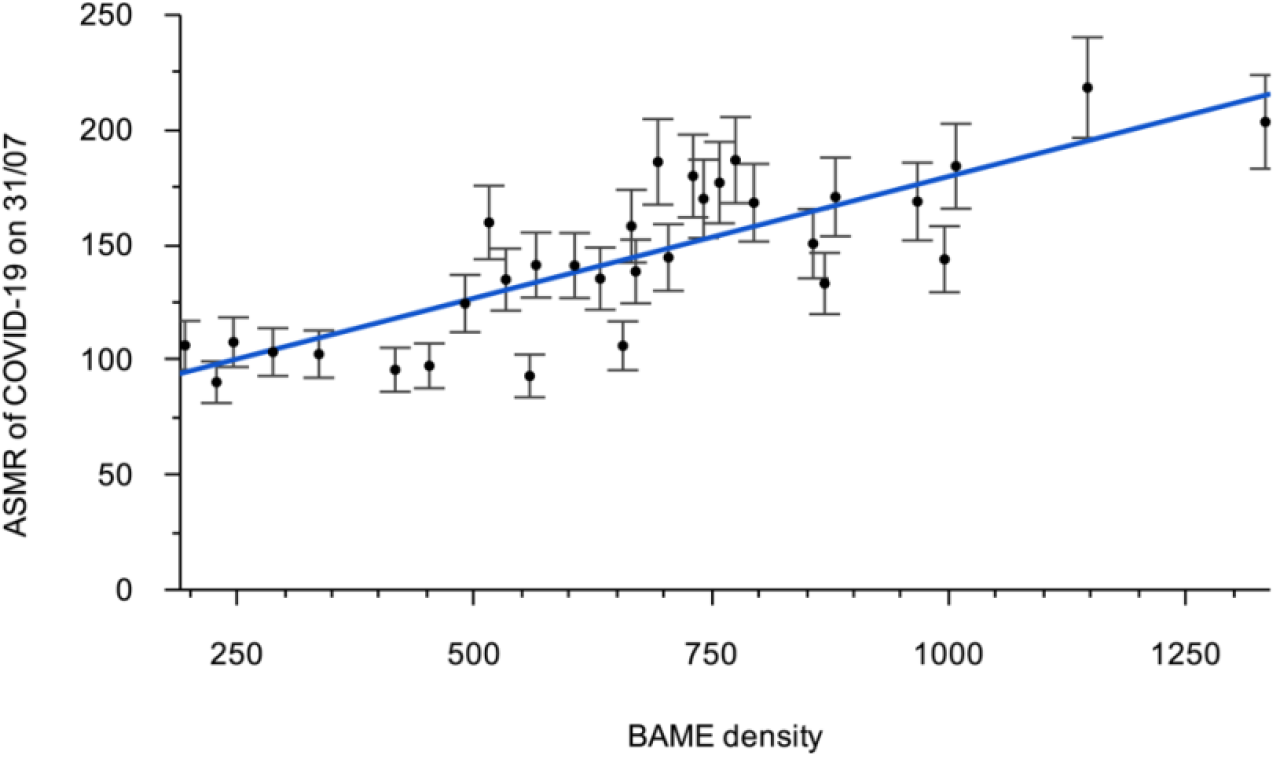
Linear regression of age-standardised COVID-19 mortality rates and BAME population density across London. Although no causal relationship can be inferred, the direction of the relationship is strongly positive.

Fig.4 suggests a linear relationship between COVID-19 mortality and the density of BAME communities. Although no causal relationship can be inferred, there is a clear positive correlation.

In order to qualitatively assess how structural racism could affect BAME populations during a pandemic, we mapped the following three datasets:

1. Age-standardised COVID-19 mortality per 100,000 population (April and July 2020 data)
2. BAME population densities per 100,000 population (2011 census)
3. IMD 2019 weighted average score per borough

We mapped A and B (Fig.5), B and C (Fig.6), A and C (Fig.7), and A, B and C (Fig.8). The boroughs that consistently score the lowest are Newham and Brent, with Richmond-Upon-Thames scoring the highest.

## Discussion

The results of the Pearson’s correlation indicate a strong positive relationship between BAME population density and the age-standardised COVID-19 mortality rates, both at the peak and end of the first wave. Moreover, a high positive correlation was identified between deprivation level, as measured by the IMD score, and the age-standardised COVID-19 mortality rates for each borough.

These results suggest that COVID-19 mortality might have disproportionately affected BAME communities, although the COVID-19 mortality data is not separated into ethnicities. Moreover, areas that were more highly deprived, and therefore experienced higher psychosocial stressors, were found to correspond to high mortality rates from the virus. This warrants further investigation to understand how exactly biological inequity played a role in the disparities of COVID-19 at a community level. It would also be of benefit to compare different communities to understand the intricate lived experience drivers which might affect susceptibility to the virus.

The results of the mapping provide a qualitative visual way of investigating the relationship between structural racism and COVID-19 mortality rates. Figures 5-8 suggest a systematic application of structural racism, which could possibly be the cause of the high mortality rates due to COVID-19 experienced in certain London boroughs. The maps reveal a pattern of BAME communities being subjected to high levels of deprivation across all domains including housing, environment, and health access.

**Figure 5.**
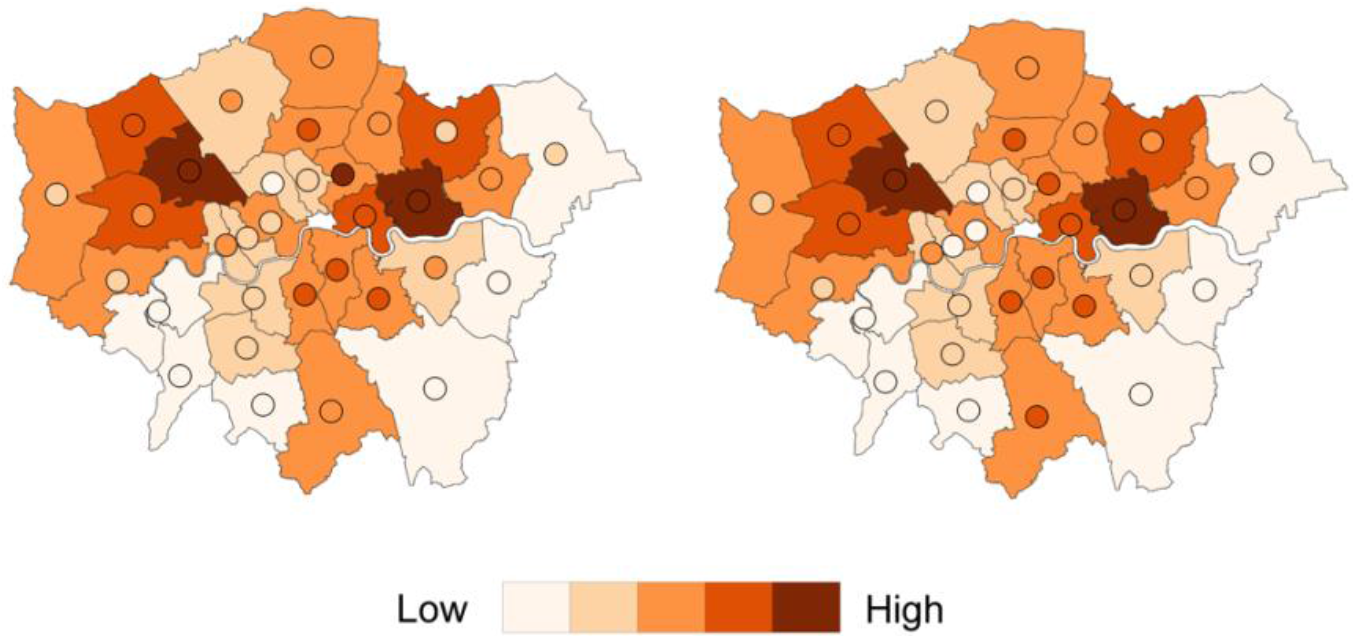
BAME population density (map) overlayed with age-standardised COVID-19 deaths (circles) by end of April (left) and July (right) 2020 in London. Dark orange denotes the areas with high values in either BAME density or COVID-19 deaths. Where both the circles and the borough are dark orange, it indicates a high positive correlation of BAME density with COVID-19 mortality. Newham and Brent score the highest.

When proposing that deprivation and structural racism may be a cause for the high number of deaths by COVID-19 among BAME communities, the results here have provided some correlations while the background offers possibilities for causality. This discussion combines the two.

Figure 6 indicates structural racism in London, by overlapping data on deprivation to BAME population density. This is an important factor to note as it highlights that BAME communities are more likely to be in areas with psychosocial stressors and environmental stressors. Although the SRS data on pollution did not yield significant results for the end of the pandemic’s first peak, it did have significant correlation with deaths at the height of the first wave in April. Indeed, several studies have pointed to a negative role of air pollution in the severity of COVID-19 outcomes.^113,114^ It is important to point out that the SRS and IMD scores are positively correlated, likely because the IMD encompasses environmental deprivation as one of its domains. Due to the possible dependence of the two variables, the IMD was used throughout this study as the representative variable. Nevertheless, we predict that biological inequity (i.e. the composite of IMD and SRS) is disproportionately experienced by BAME communities in London, since both SRS and IMD were significantly correlated to BAME population density.

**Figure 6.**
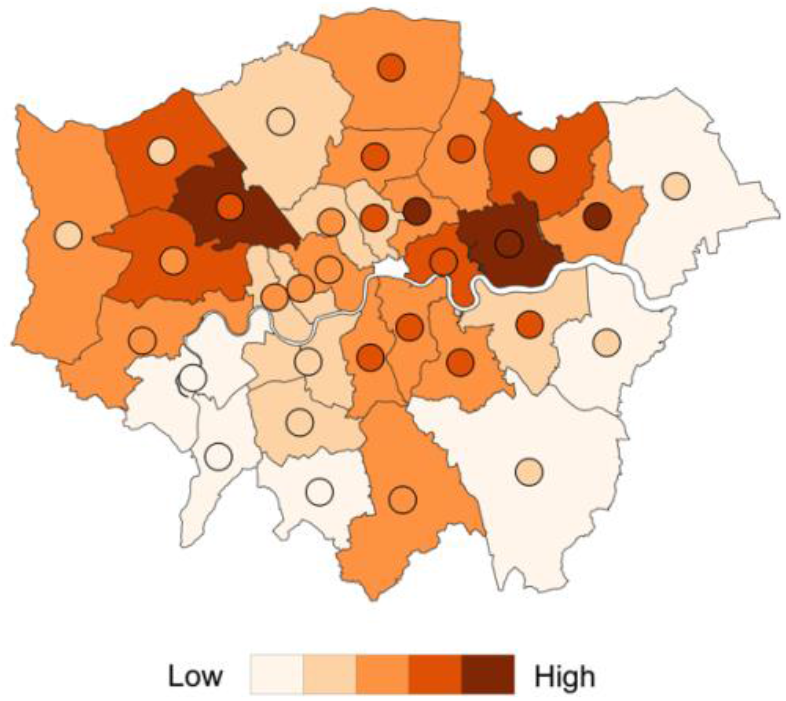
BAME population density (map) overlayed with IMD 2019 scores (circles) in London. Dark orange denotes the areas with high values in either BAME density or deprivation. Where both the circles and the borough are dark orange, it indicates a high correlation of BAME density with deprivation. Newham is the most deprived area with highest BAME density.

In terms of COVID-19 when the death rate was compared to the health risks due to deprivation (Fig. 7) there was a strong overlap. For example, Hackney and Tower Hamlets both have high COVID-19 death rates and high health risk. This indicates that identifying hotspots prior to the pandemic could have been useful for deploying extra health resources such as personal protective equipment and testing kits in those areas, as well as targeting messages to the populations including through the main languages in those locations. The maps also indicate that susceptibility though a strong factor in disease pathology is not deterministic since other factors such as jobs, emotional support through social cohesion, diet, and other health indicators are also at play. Conversely, when BAME population and IMD are compared to the COVID-19 death rate map (Fig. 8) there is an even stronger overlap, indicating that BAME communities are likely experiencing health inequities. Some possible scenarios are that BAME communities could be living in these areas for longer, could hold jobs exposing them more to further stressors, live in abodes with higher population densities or more confined spaces such as lifts, have less access to health facilities, or might shop or socialise more in person rather than online. This is worth further investigation at a population level.

**Figure 7.**
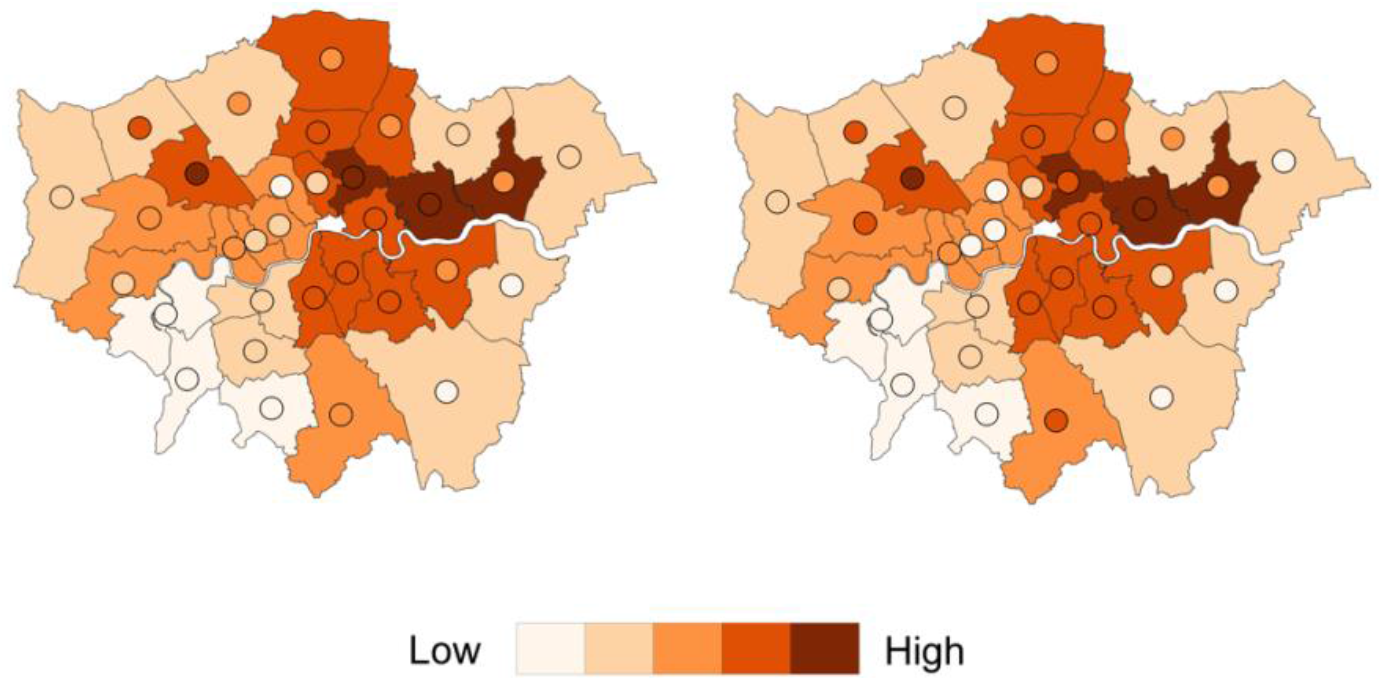
IMD 2019 scores (map) overlayed with age-standardised COVID-19 deaths (circles) by end of April (left) and July (right) 2020 in London. Dark orange denotes the areas with high values in either COVID-19 mortality or deprivation. Where both the circles and the borough are dark orange, it indicates a high correlation of COVID-19 mortality with deprivation. North-east London is most affected.

**Figure 8.**
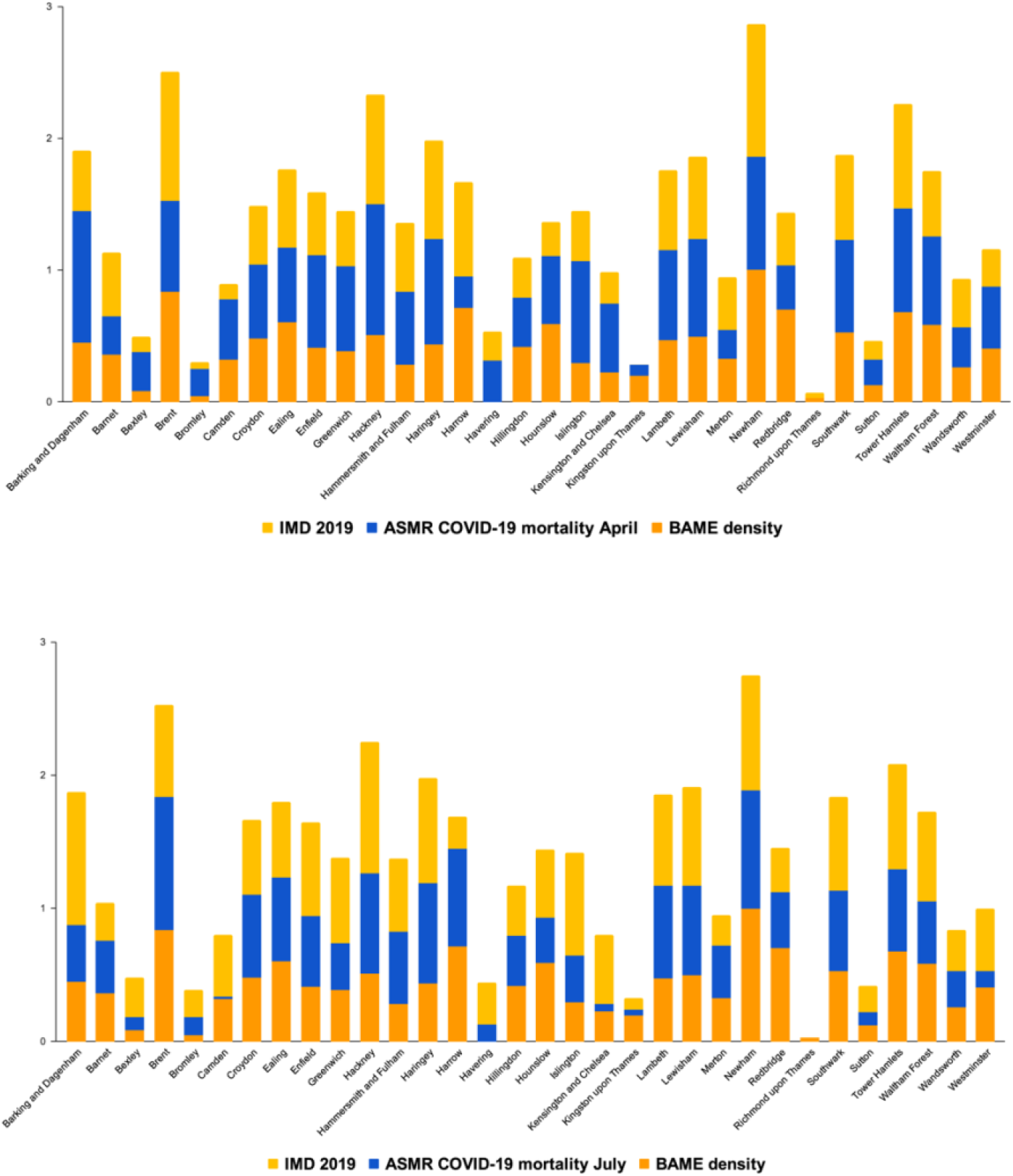
Stacked histogram of IMD 2019 normalised scores, normalised BAME population density and normalised age-standardised COVID-19 deaths by end of April (top) and July (bottom) 2020 in London. Newham scores the highest in all three measures, followed by Brent.

The data maps point to how structural racism plays a role in the story of COVID-19. It manifests in BAME communities tending to live in areas of higher environmental pollutants and deprivation. It is possible this has made these communities more susceptible to chronic disorders and so they display higher allostatic load levels, which have heightened both the risk of contracting COVID-19 and mortality from it.

Biological equity and health equity in urban systems does not mean returning to the previous norm, or merely recovering from the resultant damage. Rather, it is an adaptive, flexible, iterative process where learnings from previous failings are acknowledged in order to address the underlying structural conditions which resulted in the exposed health inequities (i.e. susceptibility and vulnerability),^115^ thereby leading to the pandemic as a disaster. To achieve positive action on biological equity, including contextual grounding, it is vital that local level involvement and initiatives are supported.^5^ This means co-designing interventions combining technical and socio-political perspectives and aspects, especially poverty-based and discrimination-based vulnerability.^116^

For example, a case study addressed the higher susceptibility to air pollution in a predominantly low-SES BAME community in London by advocating for stricter industry protocols and increased community empowerment.^117^ This example demonstrates Centric Labs’ concept of ‘biological inequity’ encompassing both technical and socio-political aspects by acknowledging the intersection between the built environment, poverty, and structural racism, and how this impacts health through the stress-response system. Relating this back to COVID-19, this means designing interventions that reduce biological inequity for improving health. Purely medical interventions will not suffice, which is the importance of the planetary health ethos. Instead, structural changes to urban systems are needed factoring in and tackling environmental racism and inequities which, in turn, improves health at the individual level.

What has been learned from this London data study is that structural racism plays a role in biological inequity, which in turn plays a role in COVID-19 susceptibility. It would be reasonable to predict that a similar phenomenon would be the case in major cities across the Americas and Europe where racism is vast.

## Data Availability

The data in this manuscript has been obtained from the UK government open source database.

## Authors Contributions

AC conceived the study and wrote the manuscript with help from all other authors. EH coined the term ‘biological inequity’ and its meaning. DOA and SA did the analysis on the London data.

## References

1. COVID-19 Report. ICNARC https://www.icnarc.org/Our-Audit/Audits/Cmp/Reports.

2. Sequist Thomas D. The Disproportionate Impact of Covid-19 on Communities of Color. Catalyst non-issue content 1, (2020).

3. Kakol, M., Upson, D. & Sood, A. Susceptibility of Southwestern American Indian Tribes to Coronavirus Disease 2019 (COVID-19). J. Rural Health (2020) doi:10.1111/jrh.12451.

4. Yancy, C. W. COVID-19 and African Americans. JAMA 323, 1891–1892 (2020).

5. Kelman, I. Relocalising disaster risk reduction for urban resilience. (2008) doi:10.1680/UDAP.2008.161.4.197.

6. Kelman, I. COVID-19: what is the disaster? Soc Anthropol (2020) doi:10.1111/1469-8676.12890.

7. World Health Organization. Essential steps for developing or updating a national pandemic influenza preparedness plan. (2018).

8. Swindells, M. Board Paper - NHS England. (2017).

9. Burkle, F. M. & Devereaux, A. V. 50 States or 50 Countries: What Did We Miss and What Do We Do Now? Prehosp. Disaster Med. 35, 353–357 (2020).

10. Pollock, A. M., Roderick, P., Cheng, K. K. & Pankhania, B. Covid-19: why is the UK government ignoring WHO’s advice? BMJ 368, m1284 (2020).

11. Hunter, D. J. Covid-19 and the Stiff Upper Lip - The Pandemic Response in the United Kingdom. N. Engl. J. Med. 382, e31 (2020).

12. Clair, M. & Denis, J. S. Racism, Sociology of. in International Encyclopedia of the Social & Behavioral Sciences (Second Edition) (ed. Wright, J. D.) 857–863 (Elsevier, 2015).

13. Pye, S., King, K. & Sturman, J. Air Quality and Social Deprivation in the UK: an environmental inequalities analysis. (2006).

14. Reese, A. M. Race, Self-Reliance, and Food Access in Washington, D.C. (University of North Carolina Press, 2019).

15. Sundberg, M. A. et al.. Implementation of the Navajo fruit and vegetable prescription programme to improve access to healthy foods in a rural food desert. Public Health Nutr. 23, 2199–2210 (2020).

16. Bowie, P. The English Indices of Deprivation 2019 (IoD2019). GOV. UK (2019).

17. Takahashi, B., Adams, E. A. & Nissen, J. The Flint water crisis: local reporting, community attachment, and environmental justice. Local Environ. 25, 365–380 (2020).

18. Colmer, J., Hardman, I., Shimshack, J. & Voorheis, J. Disparities in PM2.5 air pollution in the United States. Science 369, 575–578 (2020).

19. Bashir, S. A. Home is where the harm is: inadequate housing as a public health crisis. Am. J. Public Health 92, 733–738 (2002).

20. Charmandari, E., Tsigos, C. & Chrousos, G. Endocrinology of the stress response. Annu. Rev. Physiol. 67, 259–284 (2005).

21. McEwen, B. S. The neurobiology of stress: from serendipity to clinical relevance. Brain Research Interactive 886, 172–189 (2000).

22. Herman, J. P. et al.. Regulation of the Hypothalamic-Pituitary-Adrenocortical Stress Response. Compr. Physiol. 6, 603–621 (2016).

23. Smith, S. M. & Vale, W. W. The role of the hypothalamic-pituitary-adrenal axis in neuroendocrine responses to stress. Dialogues Clin. Neurosci. 8, 383–395 (2006).

24. Camargo, A., Hossain, E., Aliko, S., Akinola-Odusola, D. & Artus, J. Neuroscience, urban regeneration and urban health. Journal of Urban Regeneration & Renewal 13, 280–289 (2020).

25. WHO. Constitution of the World Health Organization. (2006).

26. McCartney, G., Popham, F., McMaster, R. & Cumbers, A. Defining health and health inequalities. Public Health 172, 22–30 (2019).

27. Siegrist, J. & Marmot, M. Health inequalities and the psychosocial environment-two scientific challenges. Soc. Sci. Med. 58, 1463–1473 (2004).

28. Egan, M., Tannahill, C., Petticrew, M. & Thomas, S. Psychosocial risk factors in home and community settings and their associations with population health and health inequalities: a systematic meta-review. BMC Public Health 8, 239 (2008).

29. Gochfeld, M. & Burger, J. Disproportionate exposures in environmental justice and other populations:the importance of outliers. Am. J. Public Health 101 Suppl 1, S53–63 (2011).

30. Nurse, J. & Edmondson-Jones, P. A framework for the delivery of public health: an ecological approach. J. Epidemiol. Community Health 61, 555–558 (2007).

31. Nurse, J., Basher, D., Bone, A. & Bird, W. An ecological approach to promoting population mental health and well-being – A response to the challenge of climate change. Royal Society for Public Health (2010).

32. Dubowsky, S. D., Suh, H., Schwartz, J., Coull, B. A. & Gold, D. R. Diabetes, obesity, and hypertension may enhance associations between air pollution and markers of systemic inflammation.Environ. Health Perspect. 114, 992–998 (2006).

33. van der Valk, E. S., Savas, M. & van Rossum, E. F. C. Stress and Obesity: Are There More Susceptible Individuals? Curr. Obes. Rep. 7, 193–203 (2018).

34. Grimm, N. B., Grove, J. G., Pickett, S. T. A. & Redman, C. L. Integrated Approaches to Long-Term Studies of Urban Ecological SystemsUrban ecological systems present multiple challenges to ecologists—pervasive human impact and extreme heterogeneity of cities, and the need to integrate social and ecological approaches, concepts, and theory. Bioscience 50, 571–584 (2000).

35. International Organization for Migration. World Migration Report 2020.https://publications.iom.int/books/world-migration-report-2020 (2020).

36. Mills, J. G. et al.. Relating Urban Biodiversity to Human Health With the ‘Holobiont’ Concept. Front. Microbiol. 10, 550 (2019).

37. Nadybal, S. M., Collins, T. W. & Grineski, S. E. Light pollution inequities in the continental United States: A distributive environmental justice analysis. Environ. Res. 189, 109959 (2020).

38. L., H. S. et al. In the shade of affluence: the inequitable distribution of the urban heat island. in Equity and the Environment (eds. Wilkinson, R. C. & Freudenburg, W. R.) vol. 15 173–202 (Emerald Group Publishing Limited, 2007).

39. Li, H. & Wei, Y. D. Spatial inequality of housing value changes since the financial crisis. Appl. Geogr. 115, 102141 (2020).

40. Jonkman, S. N. & Kelman, I. An analysis of the causes and circumstances of flood disaster deaths. Disasters 29, 75–97 (2005).

41. Jorgenson, A. K. et al.. Power, proximity, and physiology: does income inequality and racial composition amplify the impacts of air pollution on life expectancy in the United States? Environ. Res. Lett. 15, 024013 (2020).

42. Zhang, W., Villarini, G., Vecchi, G. A. & Smith, J. A. Urbanization exacerbated the rainfall and flooding caused by hurricane Harvey in Houston. Nature (2018).

43. Houston, D., Wu, J., Ong, P. & Winer, A. Structural Disparities of Urban Traffic in Southern California: Implications for Vehicle-Related Air Pollution Exposure in Minority and High-Poverty Neighborhoods. J. Urban Aff. 26, 565–592 (2004).

44. DiPrete, T. A. The Impact of Inequality on Intergenerational Mobility. Annu. Rev. Sociol. (2020)doi:10.1146/annurev-soc-121919-054814.

45. Bailey, Z. D. et al.. Structural racism and health inequities in the USA: evidence and interventions. Lancet 389, 1453–1463 (2017).

46. Brulle, R. J. & Pellow, D. N. Environmental justice: human health and environmental inequalities. Annu. Rev. Public Health 27, 103–124 (2006).

47. Nazroo, J. & Becares, L. Evidence for ethnic inequalities in mortality related to COVID-19 infections: Findings from an ecological analysis of England and Wales. Epidemiology (2020)doi:10.1101/2020.06.08.20125153.

48. Health of People, Places and Planet: Reflections based on Tony McMichael’s four decades of contribution to epidemiological understanding. (ANU Press, 2015).

49. Marmot, M. Fair Society, Healthy Lives. Strategic Review of Health Inequalities in England.

50. Münzel, T. & Daiber, A. Environmental Stressors and Their Impact on Health and Disease with Focus on Oxidative Stress. Antioxid. Redox Signal. 28, 735–740 (2018).

51. Guilliams, T. G. & Edwards, L. Chronic Stress and the HPA Axis: Clinical Assessment and Therapeutic Considerations. The Standard (2010).

52. Thomson, E. M. Air Pollution, Stress, and Allostatic Load: Linking Systemic and Central Nervous System Impacts. J. Alzheimers. Dis. 69, 597–614 (2019).

53. Gunnar, M. & Quevedo, K. The neurobiology of stress and development. Annu. Rev. Psychol. 58, 145–173 (2007).

54. Cool, J. & Zappetti, D. Allostasis: The Normal Stress Response and the Reason that Stress Exists. Nature (2019).

55. Freeman, B. M. Physiological basis of stress. Proc. R. Soc. Med. 68, 427–429 (1975).

56. Clougherty, J. E. & Kubzansky, L. D. A framework for examining social stress and susceptibility to air pollution in respiratory health. Environ. Health Perspect. 117, 1351–1358 (2009).

57. McEwen, B. S. & Wingfield, J. C. What is in a name? Integrating homeostasis, allostasis and stress. Hormones and behavior vol. 57 105–111 (2010).

58. Sterling, P. & Eyer, J. Allostasis: A New Paradigm to Explain Arousal Pathology. Handbook of Life Stress, Cognition and Health (1988).

59. McEwen, B. S. Stressed or stressed out: what is the difference? J. Psychiatry Neurosci. 30, 315–318 (2005).

60. McEwen, B. S. & Tucker, P. Critical biological pathways for chronic psychosocial stress and research opportunities to advance the consideration of stress in chemical risk assessment. Am. J. Public Health 101 Suppl 1, S131–9 (2011).

61. Baptiste, D. et al.. Racial discrimination in health care: An ‘us’ problem. J. Clin. Nurs. 29, 4415–4417 (2020).

62. Canady, V. A. Bill would improve MH service access for BIPOC communities. Mental Health Weekly 30, 3–5 (2020).

63. Stroud, P. Measuring Poverty 2020. https://socialmetricscommission.org.uk/wp-content/uploads/2020/06/Measuring-Poverty-2020-Web.pdf (2020).

64. Steckler, T. Chapter 1.2 - The neuropsychology of stress. in Techniques in the Behavioral and Neural Sciences (eds. Steckler, T., Kalin, N. H. & Reul, J. M. H. M.) vol. 15 25–42 (Elsevier, 2005).

65. Gu, J. et al.. Ambient air pollution and cause-specific risk of hospital admission in China: A nationwide time-series study. PLoS Med. 17, e1003188 (2020).

66. Ranveer, A. C., Pawar, P. & Latake, P. T. Odour Pollution and Its Measurement. 3, 221–229 (2015).

67. Kumar, D. & Kumar, D. Chapter 13 - Abatement of Noise Pollution. in Sustainable Management of Coal Preparation (eds. Kumar, D. & Kumar, D.) 279–291 (Woodhead Publishing, 2018).

68. Günay, O., Yarar, O. & Sarihan, M. Determination of Environmental Noise Contamination. Acta Phys. Pol. A 137, 574–578 (2020).

69. Auger, N., Duplaix, M., Bilodeau-Bertrand, M., Lo, E. & Smargiassi, A. Environmental noise pollution and risk of preeclampsia. Environ. Pollut. 239, 599–606 (2018).

70. Cinzano, P. & Falchi, F. Quantifying light pollution. J. Quant. Spectrosc. Radiat. Transf. 139, 13–20(2014).

71. Hoffman, J. S., Shandas, V. & Pendleton, N. The Effects of Historical Housing Policies on Resident Exposure to Intra-Urban Heat: A Study of 108 US Urban Areas. Climate 8, 12 (2020).

72. Public Health England. Beyond the data: Understanding the impact of COVID-19 on BAME groups.https://assets.publishing.service.gov.uk/government/uploads/system/uploads/attachment_data/file/892376/COVID_stakeholder_engagement_synthesis_beyond_the_data.pdf (2020).

73. Jacobs, D. E. Environmental health disparities in housing. Am. J. Public Health 101 Suppl 1, S115–22 (2011).

74. Gulliver, K. Racial discrimination in UK housing has a long history and deep roots. http://eprints.lse.ac.uk/85294/1/politicsandpolicy-racial-discrimination-in.pdf (2017).

75. Bednar, D. J. & Reames, T. G. Recognition of and response to energy poverty in the United States. Nature Energy 5, 432–439 (2020).

76. Longhurst, N. & Hargreaves, T. Emotions and fuel poverty: The lived experience of social housing tenants in the United Kingdom. Energy Research & Social Science 56, 101207 (2019).

77. Ponnet, K. Financial stress, parent functioning and adolescent problem behavior: an actor-partner interdependence approach to family stress processes in low-, middle-, and high-income families. J.Youth Adolesc. 43, 1752–1769 (2014).

78. Matheson, K., Foster, M. D., Bombay, A., McQuaid, R. J. & Anisman, H. Traumatic Experiences,Perceived Discrimination, and Psychological Distress Among Members of Various Socially Marginalized Groups. Front. Psychol. 10, 416 (2019).

79. Sarche, M. & Spicer, P. Poverty and health disparities for American Indian and Alaska Native children: current knowledge and future prospects. Ann. N. Y. Acad. Sci. 1136, 126–136 (2008).

80. Szczepura, A. Access to health care for ethnic minority populations. Postgrad. Med. J. 81, 141–147 (2005).

81. Umlauf, M. G., Bolland, A. C., Bolland, K. A., Tomek, S. & Bolland, J. M. The effects of age,gender, hopelessness, and exposure to violence on sleep disorder symptoms and daytime sleepiness among adolescents in impoverished neighborhoods. J. Youth Adolesc. 44, 518–542 (2015).

82. Farrell, A. D. & Sullivan, T. N. Impact of witnessing violence on growth curves for problem behaviors among early adolescents in urban and rural settings. J. Community Psychol. 32, 505–525(2004).

83. Skybo, T. Witnessing violence: biopsychosocial impact on children. Pediatr. Nurs. 31, 263–270 (2005).

84. Mc Curdy, K., Gorman, K. S. & Metallinos-Katsaras, E. From Poverty to Food Insecurity and Child Overweight: A Family Stress Approach.

85. Martin, M. S., Maddocks, E., Chen, Y., Gilman, S. E. & Colman, I. Food insecurity and mental illness: disproportionate impacts in the context of perceived stress and social isolation. Public Health132, 86–91 (2016).

86. Swiersz, S. Sea-Level Rise and Climate Justice for Native Americans and Indigenous Peoples: An Analysis of the United States’ Response and Responsibilities.https://stars.library.ucf.edu/honorstheses/805 (2020).

87. Cianconi, P., Betrò, S. & Janiri, L. The Impact of Climate Change on Mental Health: A Systematic Descriptive Review. Front. Psychiatry 11, 74 (2020).

88. Lee, J.-Y., Kim, S.-W. & Kim, J.-M. The Impact of Community Disaster Trauma: A Focus on Emerging Research of PTSD and Other Mental Health Outcomes. Chonnam Med. J. 56, 99–107 (2020).

89. Watts, N., Amann, M., AyebKarlsson, S., Belesova, K., Bouley, T., Boykoff, M., Byass, P., Cai, W.,CampbellLendrum, D., Chambers, J., Cox, P. M., Daly, M., Dasandi, N., Davies, M., Depledge, M.,Depoux, A., DominguezSalas, P., Drummond, P., Ekins, P., Flahault, A., Frumkin, H., Georgeson, L.,Ghanei, M., Grace, D., Graham, H., Grojsman, R., Haines, A., Hamilton, I., Hartinger, S., Johnson, A., Kelman, I., Kiesewetter, G., Kniveton, D., Liang, L., Lott, M., Lowe, R., Mace, G., Odhiambo Sewe, M., Maslin, M., Mikhaylov, S., Milner, J., Latifi, A. M., MoradiLakeh, M., Morrissey, K.,Murray, K., Neville, T., Nilsson, M., Oreszczyn, T., Owfi, F., Pencheon, D., Pye, S., Rabbaniha, M.,Robinson, E., Rocklöv, J., Schütte, S., ShumakeGuillemot, J., Steinbach, R., Tabatabaei, M., Wheeler, N., Wilkinson, P., Gong, P., Montgomery, H. and Costello, A. The Lancet Countdown on health and climate change: from 25 years of inaction to a global transformation for public health. The Lancet (2018) doi:10.1016/S01406736(17)324649.

90. Munro, A. et al.. Effect of evacuation and displacement on the association between flooding and mental health outcomes: a cross-sectional analysis of UK survey data. Lancet Planet Health 1, e134–e141 (2017).

91. Campagna, G. Linking crowding, housing inadequacy, and perceived housing stress. J. Environ. Psychol. 45, 252–266 (2016).

92. Mangrio, E. & Zdravkovic, S. Crowded living and its association with mental ill-health among recently-arrived migrants in Sweden: a quantitative study. BMC Res. Notes 11, 609 (2018).

93. Adger, W. N., Safra de Campos, R., Siddiqui, T. & Szaboova, L. Commentary: Inequality, precarity and sustainable ecosystems as elements of urban resilience. Urban Stud. 57, 1588–1595 (2020).

94. Wiesel, I. & Liu, F. Conceptualising modes of redistribution in public urban infrastructure. Urban Stud. 0042098020913188 (2020).

95. Lopez, B., Kennedy, C. & McPhearson, T. Parks are Critical Urban Infrastructure: Perception and Use of Urban Green Spaces in NYC During COVID-19. (2020)doi:10.20944/preprints202008.0620.v1.

96. Colder, C. R., Mott, J., Levy, S. & Flay, B. The relation of perceived neighborhood danger to childhood aggression: a test of mediating mechanisms. Am. J. Community Psychol. 28, 83–103 (2000).

97. Ross, C. E. & Mirowsky, J. Neighborhood disadvantage, disorder, and health. J. Health Soc. Behav. 42, 258–276 (2001).

98. Collins, K., Connors, K., Davis, S., Donohue, A., Gardner, S., Goldblatt, E., Hayward, A., Kiser, L., Strieder, F. Thompson, E. Understanding the impact of trauma and urban poverty on family systems:Risks, resilience and interventions. https://www.nctsn.org/sites/default/files/resources/resource-guide/understanding_impact_trauma_urban_poverty_family_systems.pdf (2010).

99. Douglas, I. Urban ecology and urban ecosystems: understanding the links to human health and well-being. Current Opinion in Environmental Sustainability 4, 385–392 (2012).

100. Danese, A. & McEwen, B. S. Adverse childhood experiences, allostasis, allostatic load, and age-related disease. Physiol. Behav. 106, 29–39 (2012).

101. Aliko, S. & Camargo, A. Air Pollution, Susceptibility & COVID-19 Learnings.https://www.thecentriclab.com/air-pollution-susceptibility-covid-19-learnings (2020).

102. Camargo, A. PTSD Prevalence in Impoverished Urban Environments.https://www.thecentriclab.com/ptsd-cities (2019).

103. Alam, M. R., Kabir, M. R. & Reza, S. Comorbidities might be a risk factor for the incidence of COVID-19: Evidence from a web-based survey of 780,961 participants. Public and Global Health (2020) doi:10.1101/2020.06.22.20137422.

104. Lighter, J. et al.. Obesity in Patients Younger Than 60 Years Is a Risk Factor for COVID-19 Hospital Admission. Clin. Infect. Dis. 71, 896–897 (2020).

105. Dhabhar, F. S. Enhancing versus Suppressive Effects of Stress on Immune Function: Implications for Immunoprotection versus Immunopathology. Allergy Asthma Clin. Immunol. 4, 2–11 (2008).

106. Cohen, S., Tyrrell, D. A. & Smith, A. P. Psychological stress and susceptibility to the common cold. N. Engl. J. Med. 325, 606–612 (1991).

107. Brody, G. H., Lei, M.-K., Chen, E. & Miller, G. E. Neighborhood poverty and allostatic load in African American youth. Pediatrics 134, e1362–8 (2014).

108. The London Plan.https://www.london.gov.uk/sites/default/files/the_london_plan_2016_jan_2017_fix.pdf (2016).

109. London Borough Profiles and Atlas. https://data.london.gov.uk/dataset/london-borough-profiles.

110. Ministry of Housing, Communities & Local Government. English indices of deprivation 2015. GOV.UK https://www.gov.uk/government/statistics/english-indices-of-deprivation-2015 (2015).

111. Home - Office for National Statistics. https://www.ons.gov.uk/.

112. Ministry of Housing, Communities & Local Government. Lower Layer Super Output Areas (LSOAs).(2020).

113. Srivastava, A. COVID-19 and air pollution and meteorology-an intricate relationship: A review. Chemosphere 263, 128297 (2021).

114. Razzaq, A., Sharif, A., Aziz, N., Irfan, M. & Jermsittiparsert, K. Asymmetric link between environmental pollution and COVID-19 in the top ten affected states of US: A novel estimations from quantile-on-quantile approach. Environ. Res. 191, 110189 (2020).

115. Kelman, I., Gaillard, J. C. & Mercer, J. Climate Change’s Role in Disaster Risk Reduction’s Future:Beyond Vulnerability and Resilience. International Journal of Disaster Risk Science 6, 21–27 (2015).

116. Weichselgartner, J. & Kelman, I. Challenges and opportunities for building urban resilience. ITU A|Z (2014).

117. Air Pollution and Health in Southall. https://static1.squarespace.com/static/57a5a729414fb58fa3e0e0a6/t/5ebd02d24fa06d4c4cd00b20/1589445337817/Southall+Report.pdf (2020).

